# Circulating serum miRNAs predict response to platinum chemotherapy in high-grade serous ovarian cancer

**DOI:** 10.1101/2024.02.04.24302321

**Authors:** Kazuhiro Suzuki, Akira Yokoi, Juntaro Matsuzaki, Kosuke Yoshida, Yusuke Yamamoto, Tomoyasu Kato, Mitsuya Ishikawa, Takahiro Ochiya, Hiroaki Kajiyama

## Abstract

**Background:** Platinum chemotherapy is the cornerstone of treatment for high-grade serous ovarian cancer (HGSOC); however, validated biomarkers that can accurately predict platinum response are lacking. Based on their roles in the underlying pathophysiology, circulating microRNAs are potential, noninvasive biomarkers in cancer. In the present study, we aimed to evaluate the circulating miRNA profiles of patients with HGSOC and to assess their potential utility as biomarkers to predict platinum response.

**Methods:** Pretreatment serum samples collected from patients who received platinum chemotherapy for stage III–IV HGSOC between 2008 and 2016 were analyzed using miRNA microarray. LASSO logistic regression analysis was used to construct predictive models for treatment-free interval of platinum (TFIp).

**Results:** The median follow-up was 54.6 (range, 3.5–144.1) months. The comprehensive analysis of 2,588 miRNAs was performed in serum samples of 153 eligible patients, and predictive models were constructed using a combination of circulating miRNAs with an area under the receiver operating characteristic curve of 0.944 for TFIp > 1 month, 0.637 for TFIp ≥ 6 months, 0.705 for TFIp ≥ 12 months, and 0.938 for TFIp ≥ 36 months. Each predictive model provided a significant TFIp classification (*p* = 0.001 in TFIp >1 month, *p* = 0.013 in TFIp ≥ 6 months, *p* < 0.001 in TFIp ≥ 12 months, and *p* < 0.001 in TFIp ≥ 36 months).

**Conclusion:** Circulating miRNA profiles has potential utility in predicting platinum response in patients with HGSOC and can aid clinicians in choosing appropriate treatment strategies.

## Introduction

According to the GLOBOCAN estimates, 313,959 new ovarian cancer cases and 207,252 ovarian cancer-related deaths occurred worldwide in 2020 [1]. High-grade serous ovarian cancer (HGSOC), the main epithelial ovarian cancer subtype [2], is already in advanced stage at diagnosis in most cases; therefore, the 5-year survival rate of advanced-stage HGSOC is approximately 30% [3]. The standard treatment strategy for HGSOC is primary debulking surgery followed by chemotherapy or neoadjuvant chemotherapy with subsequent interval debulking surgery [4]. The introduction of bevacizumab and Poly(ADP-ribose) polymerase inhibitors (PARPi) has expanded the spectrum of available treatment alternatives, but consistent therapy efficacy is limited [5, 6]. Platinum chemotherapy remains the most vital therapeutic agent for the treatment of both primary and recurrent HGSOC [4].

Treatment-free interval of platinum (TFIp) is a commonly used decision criterion for platinum rechallenge [7]. Similarly, the TFIp status has been used to classify patients in clinical trials on ovarian cancer [8], [9]. However, the efficacy of platinum rechallenge was clearly demonstrated in several phase II trials, which showed that platinum chemotherapy was no longer effective patients with a TFIp of <6 months [10, 11], indicating that a TFIp of <6 months was not a consistent predictor of response to platinum chemotherapy. There are various chemotherapy options for ovarian cancer, such as platinum chemotherapy with or without the addition of bevacizumab or maintenance therapy with or without the addition of bevacizumab and PARPi. For an expected short duration of TFIp with platinum chemotherapy alone, the addition of bevacizumab or enhanced maintenance therapy should be selected with the expectation of additional therapeutic benefit. In contrast, for an expected long duration of TFIp with platinum chemotherapy alone, the patient may choose not to receive any additional therapy or maintenance therapy. Therefore, accurate prediction of TFIp is essential for treatment selection. Currently, validated biomarkers that predict response to platinum chemotherapy in clinical settings are lacking.

MicroRNAs (miRNAs) are endogenous, small, noncoding, single-stranded RNAs that regulate target gene expression and play vital roles in cancer progression [12, 13]. Importantly, miRNAs are released from cells to stably exist in body fluids by escaping RNase degradation. Circulating miRNAs in body fluids are wrapped up by extracellular vesicles, thereby intervening in cell-to-cell communication in both the original and the distant microenvironments [14–16]. We have previously demonstrated the potential role of circulating miRNAs as noninvasive diagnostic biomarkers for ovarian cancer, with high accuracy and prognostic ability found in high-grade serous carcinoma and ovarian clear cell carcinoma [17–19]. Recent studies have also reported the utility of miRNA expression profiles in predicting therapeutic response in cancer [20, 21].

In the present study, we aimed to determine the utility of circulating miRNA profiles as noninvasive biomarkers to predict platinum response in HGSOC. Thus, we investigated the association between platinum response and circulating miRNA profiles before platinum therapy by analyzing patients who did not receive bevacizumab or PARPi as first-line therapy to specifically determine platinum response. Therefore, the present study provides novel evidence on the utility of circulating miRNA profiles as noninvasive biomarkers to predict platinum response in HGSOC.

## Materials and methods

### Patient selection

This is a retrospective cohort study based on miRNA profiles identified in a previous study and on the clinical information of the patients. In a previous study, we collected 4,046 serum samples from patients with ovarian tumors and healthy controls admitted or referred to the National Cancer Center Hospital between 2008 and 2016 [17]. Data are available from the NCBI database under the accession number GSE106817. The present study included the pretreatment serum samples of 442 patients with ovarian tumors. The clinical information of patients, such as age, histologic subtype, International Federation of Gynecology and Obstetrics stage, residual tumor volume, administration of neoadjuvant or adjuvant chemotherapy, recurrence, and death, was retrospectively reviewed by accessing the database of the Department of Gynecology of the National Cancer Center Hospital (Tokyo, Japan). Patients with stage III–IV HGSOC were included after the exclusion of patients who underwent surgery or chemotherapy before serum collection and those with a history of other cancers. Follow-up was performed at least every 3 months for 2 years after the last platinum treatment. At the time of follow-up, computed tomography was performed after at least every 6 months. Tumors were evaluated using contrast-enhanced computed tomography every two cycles. The response to chemotherapy was evaluated according to the Response Evaluation Criteria in Solid Tumors 1.1.

The present study was approved by the National Cancer Center Hospital Institutional Review Board (approval no. 2015-376, 2016-29), and all participants provided written informed consent.

### miRNA expression analysis

The serum samples stored in the National Cancer Center Biobank were used for comprehensive miRNA expression analysis using the 3D-Gene miRNA labeling kit and the 3D-Gene Human miRNA Oligo Chip (Toray Industries Inc., Tokyo, Japan), which was designed to detect 2,588 miRNA sequences registered in miRBase (release 21) [22]. To that end, total RNA was extracted from 300 μL of the serum samples using the 3D-Gene RNA extraction reagent (Toray Industries Inc., Tokyo, Japan). In the present study focusing on circulating miRNAs released by ovarian cancer cells, miRNAs were selected according to the criteria described in our previous study [17]. Among a total of 2,038 miRNAs, 858 miRNAs were detected in extracellular vesicles derived from at least one ovarian cancer cell line. Among these, 210 miRNAs that were identified based on a human serum dataset were included in the final analysis [17].

### Statistical analysis

In the present study, TFIp was defined as the time interval from the last platinum treatment to the time of initial tumor progression or last contact and overall survival was defined as the time interval from initial treatment to the time of death from any cause (Supplementary Figure 1). Sensitivity, specificity, and accuracy of the circulating miRNAs in predicting TFIp were determined using receiver operating characteristic (ROC) curve analysis and the area under the ROC curve (AUC) was calculated. The ROC analysis was used to determine the TFIp with the best power, which was used to develop a predictive model for platinum response. Thus, predictive index-positive cases were considered as patients with a TFIp >X months. The sensitivity showed the predictive rate of TFIp longer than X months, and the specificity showed the predictive rate of TFIp less than X months. To calculate sensitivity, specificity, and accuracy, optimal cutoff values were set based on the maximum point of the sum of sensitivity and specificity, i.e., Youden index. The 95% confidence interval (95% CI) of the AUC was calculated and plotted in the ROC curve. Two-group comparison was performed using Welch’s *t*-test. LASSO logistic regression analysis was performed using the R software (version 4.3.0; R Foundation for Statistical Computing, http://www.R-project.org), compute.es (version 0.2-4), glmnet (version 2.0-3), hash (version 2.2.6), MASS (version 7.3-45), mutoss (version 0.1-10), and pROC (version 1.8). ROC curves were compared using the DeLong test. Pearson’s χ^2^, Welch’s *t* tests, and one-way analysis of variance were performed using SPSS Statistics (version 29; IBM Corp., Armonk, NY, USA). Kaplan–Meier curves were used for the analysis of TFIp and were compared using the generalized Wilcoxon test. A two-sided *p* value <0.05 were considered statistically significant for all analyses.

## Results

A total of 422 serum samples of patients with ovarian tumors were analyzed using the miRNA microarray to obtain the comprehensive miRNA expression profiles of patients. After the exclusion of serum samples from 262 patients with other ovarian cancer subtypes, 14 patients with stage I–II cancer, 11 patients with insufficient clinical information, and 2 patients with low-quality microarray results, the profiles of 153 patients were included in the final analysis (Figure 1).

**Figure 1.**
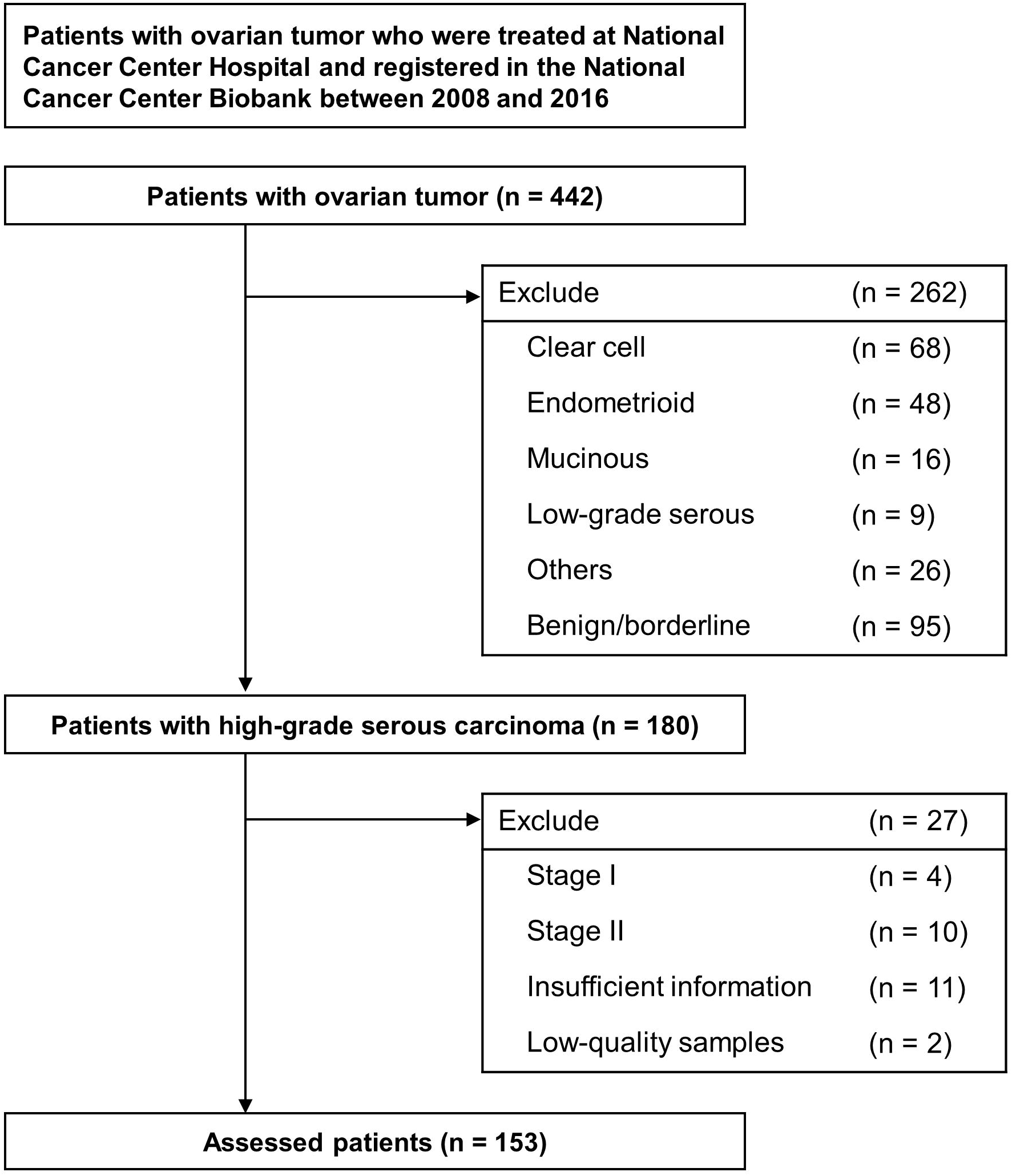
Selection of patients and study design. Flowchart of patients’ selection. We identified a total of 442 patients with ovarian tumors with preoperative serum microRNA profiles. Among them, 180 patients were diagnosed with high-grade serous ovarian carcinoma. Finally, 180 patients were assessed in this study, after excluding 27 patients with stage I–II cancer, insufficient clinical information, and low-quality samples.

The patient characteristics are summarized in Table 1. Briefly, all patients received cytoreductive surgery and adjuvant chemotherapy, which typically included carboplatin plus paclitaxel, without bevacizumab or PARPi. The median follow-up period was 54.6 (range, 3.5–144.1) months. The groups were created according to the initial TFIp, and 143, 122, 84, and 30 patients had TFIp > 1, ≥ 6, ≥ 12, and ≥ 36 months, respectively.

**Table 1.** Clinical characteristics of 153 patients with high-grade serous carcinoma grouped according to TFIp.

|  | Number of patients (%) |  |  |  |  | *p*-value |
| --- | --- | --- | --- | --- | --- | --- |
|  | Total<br>(n = 153) | TFIp > 1 months<br>(n = 143) | TFIp ≥ 6 months<br>(n = 122) | TFIp ≥ 12 months<br>(n = 84) | TFIp ≥ 36 months<br>(n = 30) |  |
| Age, years—median (range) | 60.2 (34–82) | 60.3 (34–82) | 60.5 (34–82) | 59.21 (34–82) | 56.8 (34–78) | 0.418 |
| FIGO stage |  |  |  |  |  |  |
| IIIC | 96 (62.7) | 92 (64.3) | 81 (66.4) | 56 (66.7) | 18 (60.0) | 0.981 |
| IVA | 11 (7.2) | 10 (7.0) | 9 (7.4) | 8 (9.5) | 3 (10.0) |  |
| IVB | 46 (30.1) | 41 (28.7) | 32 (26.2) | 20 (23.8) | 9 (30.0) |  |
| Neoadjuvant chemotherapy |  |  |  |  |  |  |
| Yes | 98 (62.7) | 88 (61.5) | 75 (61.5) | 47 (56.0) | 13 (43.3) | 0.255 |
| No | 55 (35.9) | 55 (38.5) | 47 (38.5) | 37 (44.0) | 17 (56.7) |  |
| Surgery |  |  |  |  |  |  |
| Complete | 9 (5.9) | 9 (6.3) | 8 (6.6) | 7 (8.3) | 6 (20.0) | 0.229 |
| Optimal | 120 (78.4) | 115 (80.4) | 99 (81.1) | 70 (83.3) | 21 (70.0) |  |
| Sub-optimal | 24 (15.7) | 19 (13.3) | 15 (12.3) | 7 (8.3) | 3 (10.0) |  |
| Recurrence |  |  |  |  |  |  |
| Yes | 132 (86.3) | 122 (85.3) | 101 (82.8) | 63 (75.0) | 10 (33.3) | < 0.001 |
| No | 21 (13.7) | 21 (14.7) | 21 (17.2) | 21 (25.0) | 20 (66.7) |  |
| TFIp, months—median (range) | 22.6 (0–130) | 24.18 (2–130) | 27.82 (6–130) | 36.49 (12–130) | 65.70 (36–130) | < 0.001 |
| PFS, months—median (range) | 28.3 (4–136) | 29.76 (5–136) | 33.38 (10–136) | 42.35 (16–136) | 72.50 (40–136) | < 0.001 |
| OS, months—median (range) | 55.8 (8–142) | 57.84 (8–142) | 64.16 (13–142) | 73.90 (22–142) | 86.03 (49–142) | < 0.001 |

Note: Data are shown as n (%) unless otherwise noted.
Abbreviation: PFS, progression-free survival. OS, overall survival.

**Table 2.**
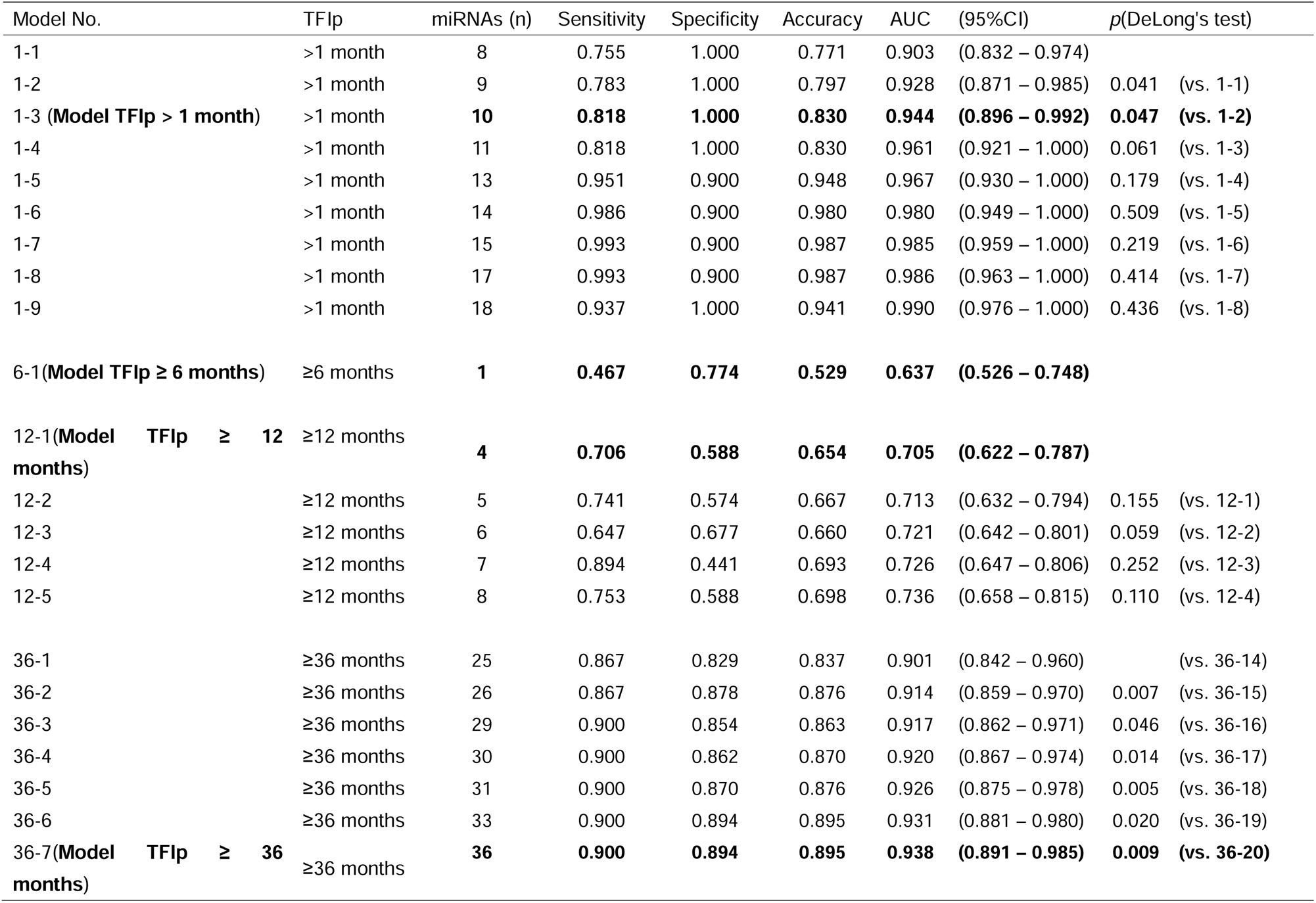
Discrimination accuracy of each diagnostic model.

### Selection of circulating miRNA candidates, and construction of predictive models

A total of 210 miRNAs selected as candidate biomarkers according to the method described in our previous report were used in combination with other prognostic factors included in Table 1 [17]. In each group, a predictive model was constructed using the LASSO logistic regression analysis. The results are summarized in Table 1, and the details of the predictive model are shown in Supplementary Table 1. Only one model was selected to predict TFIp in each group based on DeLong test. The most appropriate predictive model was expressed as follows: model TFIp > 1 month: exp Y / (1 + exp Y) | Y = (−0.43074 × miR-1228-5p) + (−0.2713 × miR-1273g-3p) + (0.2881 × miR-3917) + (−0.15449 × miR-3940-5p) + (0.03679 × miR-4708-3p) + (−1.00381 × miR-4739) + (−0.0285 × miR-486-5p) + (0.47157 × miR-498) + (0.35716 × miR-6088) + (0.09762 × miR-6872-3p) + 13.9039 and model TFIp ≥ 6 months: exp Y / (1 + exp Y) | Y = (0.04331 × miR-4708-3p) + 1.0617 and model TFIp ≥ 12 months: exp Y / (1 + exp Y) | Y = (−0.0426 × miR-3141) + (0.01861 × miR-3928-3p) + (0.0129 × miR-6766-5p) + (0.0094 × miR-7108-3p) + 0.31159, and model TFIp ≥ 12 months: exp Y / (1 + exp Y) | Y = (−0.0125433 × miR-1181) + (0.1067349 × miR-1254) + (0.0015947 × miR-1268b) + (−0.3771727 × miR-187-5p) + (0.0360527 × miR-191-5p) + (−0.0984009 × miR-3141) + (0.0233745 × miR-3195) + (−0.5606837 × miR-3197) + (0.2415774 × miR-320a) + (0.1046963 × miR-342-5p) + (0.0502296 × miR-3928-3p) + (0.0009486 × miR-422a) + (0.0050783 × miR-4419b) + (0.05179 × miR-4429) + (0.4470169 × miR-4447) + (0.0210486 × miR-4449) + (−0.9951068 × miR-4463) + (−0.0335096 × miR-4484) + (0.0152364 × miR-4515) + (0.0870314 × miR-4640-5p) + (0.2338915 × miR-4675) + (0.0864322 × miR-486-5p) + (0.5827542 × miR-557) + (0.0782236 × miR-6088) + (−0.1712249 × miR-642a-3p) + (0.1380852 × miR-6766-3p) + (−0.3991616 × miR-6799-5p) + (−0.2738122 × miR-6808-5p) + (0.0531576 × miR-6842-5p) + (−0.5248665 × miR-6858-5p) + (0.033589 × miR-7108-3p) + (−0.2512049 × miR-718) + (−0.5161003 × miR-8089) + (−0.1219326 × miR-887-3p) + (0.0626341 × miR-939-5p) + 19.9189978. The AUC of most appropriate predictive models for each model was as follows: model TFIp > 1 month: 0.944 (95% CI: 0.896–0.992) for the group with TFIp > 1 month, model TFIp ≥ 6 months: 0.637 (95% CI: 0.526–0.748) for the group with TFIp ≥ 6 months, model TFIp ≥ 12 months: 0.705 (95% CI: 0.622–0.787) for the group with TFIp ≥ 12 months, and model TFIp ≥ 36 months: 0.938 (95% CI: 0.891–0.985) for the model with TFIp ≥ 36 months (Figure 2).

**Figure 2.**
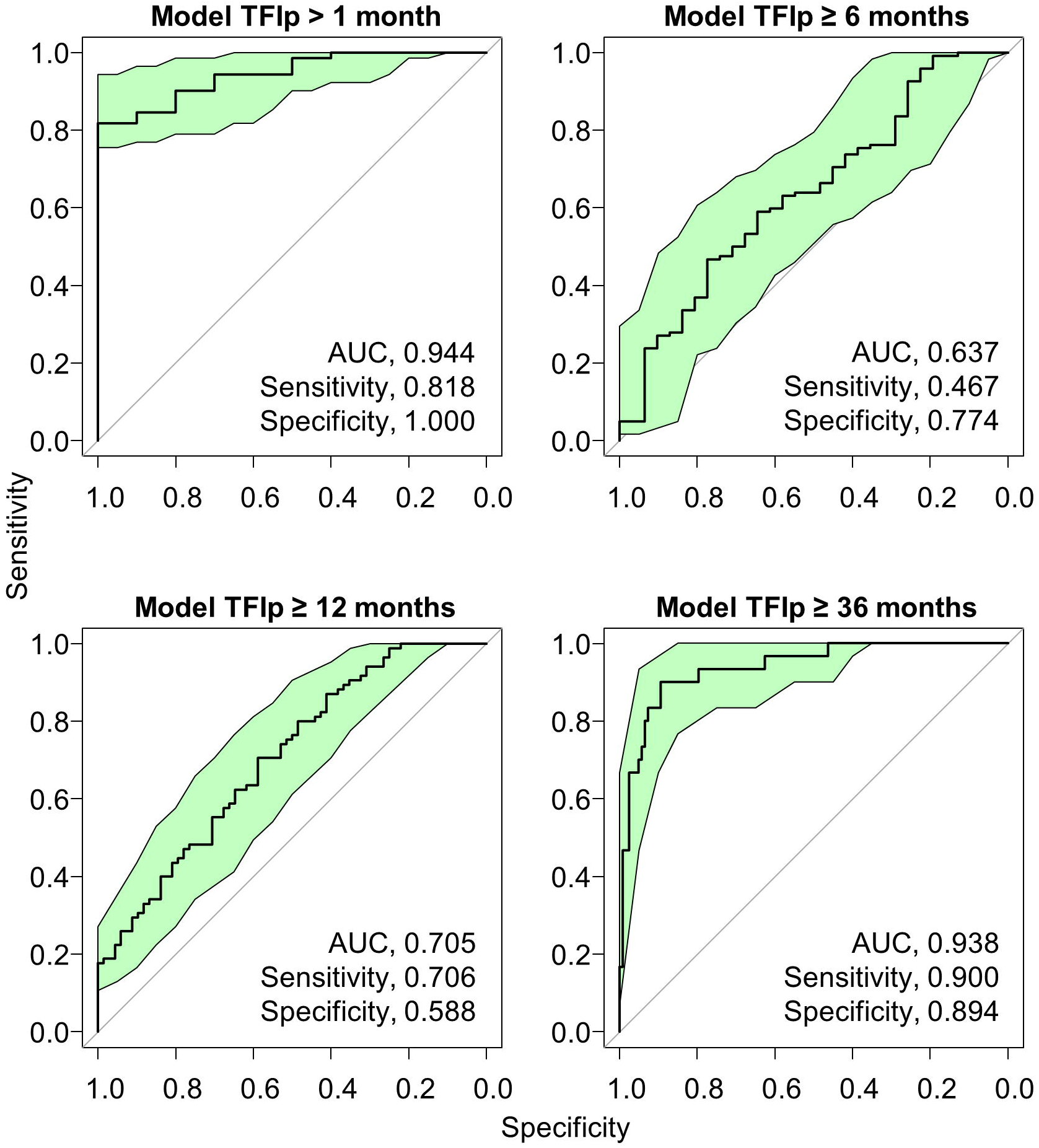
ROC curves for each Model. ROC curves for prediction of TFIp by models using LASSO logistic regression analysis. The green colored area indicates the 95% confidence interval region. The details of the predictive model are shown in Supplementary Table 1.

### Further potentials for predictive models

Each model was applied to all subgroups and AUCs were calculated. Model TFIp > 1 month: AUC of 0.683 for the group with TFIp ≥ 6 months, 0.603 for TFIp ≥ 12 months, and 0.618 for TFIp ≥ 36 months. Model TFIp ≥ 6 months: 0.631 for TFIp > 1 month, 0.570 for TFIp 12 ≥ months, and 0.563 for TFIp ≥ 36 months. Model TFIp ≥ 12 months: 0.757 for > 1 month, 0.698 for TFIp 6 ≥ months, and 0.659 for TFIp ≥ 36 months. Model TFIp ≥ 36 months: 0.624 for > 1 month, 0.641 for TFIp 6 ≥ months, and 0.645 for TFIp ≥ 12 months (Supplementary Table 2). The predictive models for each TFIp group were re-evaluated by dividing each group into those with TFIp shorter and longer than the cutoff value. As shown in Figure 3A, there was a significant difference in each group (*p* = 0.001 in model TFIp > 1 month, *p* = 0.013 in model TFIp ≥ 6 months, *p* < 0.001 in model TFIp ≥ 12 months, and *p* < 0.001 in model TFIp ≥ 36 months) (Figure 3A, Supplementary Figure 2A). Upon examining the model score transition for each patient, we observed a tendency of decreasing model score when the actual TFIp was lower than the predicted TFIp (Figure 3B).

**Figure 3.**
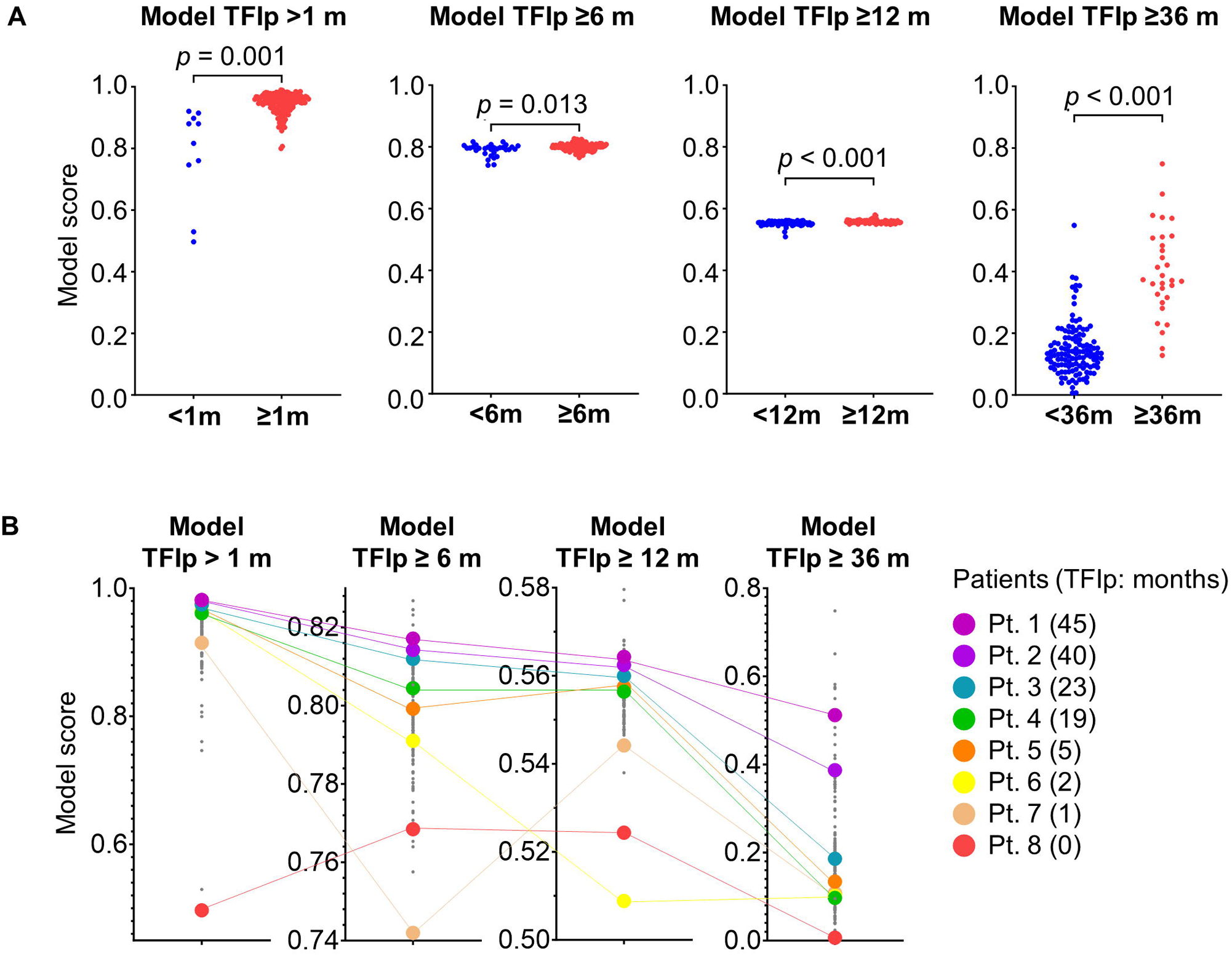
Development of the predictive TFIp models **A.** Dot plot for each Model. Patients were stratified based on the TFIp. *p* values were calculated by Welch’s *t*-test. **B.** Model values for each in the same patient. Colored values indicate representative cases.

### Reassessment of the predictive model

Pearson’s correlation coefficients were calculated between TFIp and model score for each model. TFIp was positively correlated with all models (r = 0.170 in model TFIp > 1 month, r = 0.047 in model TFIp ≥6 months, r = 0.256 in model TFIp ≥ 12 months, and r = 0.630 in model TFIp ≥ 36 months) (Supplementary Figure 2A). Kaplan–Meier curves were constructed after stratifying patients into high and low groups based on the cutoff value of each model score. The Kaplan–Meier curves indicated that patients with a high model score had significantly longer TFIp than those with a low model score (*p* = 0.006 in model TFIp > 1 month, *p* = 0.003 in model TFIp ≥ 12 months, and *p* < 0.001 in model TFIp ≥ 36 months) (Supplementary Figure 2B). Therefore, the three models have the potential to predict early recurrence, regardless of each cutoff value.

## Discussion

Accurate prediction of platinum response is fundamentally important in the clinical management of patients with HGSOC. Previous studies have explored the efficacy of platinum chemotherapy by categorizing patients as those who were platinum-sensitive and those who were platinum-resistant based on a TFIp of 6 months [23, 24]. In the present study, we evaluated the expected TFIp before platinum chemotherapy. The AUC values for model TFIp > 1 month and model TFIp ≥ 36 months were 0.944 and 0.938, respectively. The inclusion of circulating miRNA profiles in these analyses demonstrated their ability in accurately predicting TFIp.

The mechanisms underlying platinum chemotherapy resistance in patients with HGSOC have been extensively evaluated [25–28]. Platinum resistance is not straightforward and is considered a complex biologic process. Therefore, the combination of factors, rather than a single factor, is a reasonable approach to develop biomarkers. In the present study, the miRNAs selected as predictive factors were different and multiple in each model, which may reflect platinum-resistant and -sensitive pathologies. Model TFIp ≥ 12 months appeared to be versatile, with an AUC ≥0.69 for all subgroups, but model TFIp > 1 month for the group with TFIp > 1 month and model TFIp ≥ 6 months for TFIp ≥ 6 months were the most appropriate, each with an AUC of ≥0.9. The miRNA with the strongest negative impact in model TFIp > 1 month was miR-4739. With demonstrated involvement in the VEGFA/PI3K/AKT pathway, miR-4739 has been reported to play an antioncogenic role in pancreatic cancer [29]. The miRNA with the strongest impact in model TFIp ≥ 6 months was miR-4708-3p. According to miRDB, one of the targets of miR-4708-3p is interleukin 22, which has been previously reported to be involved in chemotherapy resistance in lung and breast cancers [30–32]. The miRNA with the strongest impact in model TFIp ≥ 12 months was miR-3141, which directly targets transforming growth factor-β, which has demonstrated roles in platinum response in ovarian cancer [33–35].

Currently, there is not one standard definition for therapeutic response, although biomarkers to predict therapeutic response have been investigated in ovarian cancer. In clinical trials, the typical primary endpoints are indicators such as overall and progression-free survival and response rate. Indicators that might define platinum treatment response in ovarian cancer include TFIp, response rate, and progression-free survival [24, 36, 37]. Biomarkers that can predict platinum response should be able to predict TFIp in first-line therapy because TFIp is expected. Second- or third-line chemotherapy might provide TFIp; however, recurrence and progression might still occur during platinum chemotherapy. Therefore, biomarkers predicting platinum response and recurrence in second- or third-line chemotherapy should be defined by TFIp or progression-free survival from treatment initiation for recurrence. After fourth-line therapy, the risk of recurrence and progression during platinum chemotherapy is even higher and predictive biomarkers for platinum response might be defined by the duration of continued platinum therapy. Ovarian cancer is unique as patients are repeatedly administered platinum chemotherapy, even in cases of relapse, while good response to platinum chemotherapy is expected. In the present study, we selected TFIp as an indicator of predictive biomarkers for platinum response as platinum was administered as first-line therapy. Biomarkers that can predict treatment response should be defined according to the specific line of treatment.

The present study has several limitations. First, our results were based on dataset reanalysis and validation experiments using patients with recurrent cancer could not be conducted. Second, we did not include patients who received bevacizumab or PARPi as first-line therapy; therefore, whether the predictive models can be applied to these patients remains unclear. Circulating miRNAs can be upregulated or downregulated by bevacizumab or PARPi [38, 39]. Further studies with larger, independent groups are needed to determine predictive biomarkers for response to platinum in combination with bevacizumab or PARPi. Third, although the expression of miRNAs in ovarian cancer cell lines was considered, the origin of the circulating miRNAs measured in the present study was unclear. Furthermore, the function of miRNAs in HGSOC should be evaluated in future studies.

In conclusion, we identified a circulating miRNA profile that predicted platinum response in HGSOC before platinum chemotherapy. Accurate prediction of treatment response prior to initiation can assist in determining appropriate treatment strategies, such as avoiding unnecessary treatments or choosing additional treatments. Prospective studies are warranted to evaluate circulating miRNA analysis for the translation of miRNA profiling into clinical care.

## Data Availability

the NCBI database under the accession number GSE106817

## Author Contribution

Study concept and design: Kazuhiro Suzuki., Akira Yokoi.

Provision of materials or patients: Kazuhiro Suzuki., Akira Yokoi., Kosuke Yoshida., Mitsuya Ishikawa., Tomoyasu Kato.

Analysis and interpretation of data: Kazuhiro Suzuki., Akira Yokoi., Juntaro Matsuzaki., Kosuke Yoshida.

Manuscript writing: Kazuhiro Suzuki., Akira Yokoi., Yoshida Kosuke., Juntaro Matsuzaki.

Critical review of the manuscript: All authors contributed.

Final approval of manuscript: All authors contributed.

## Acknowledgment

No funding support of any kind was received for this study.

## Supplementary Figure legends

**Supplementary Figure 1.**
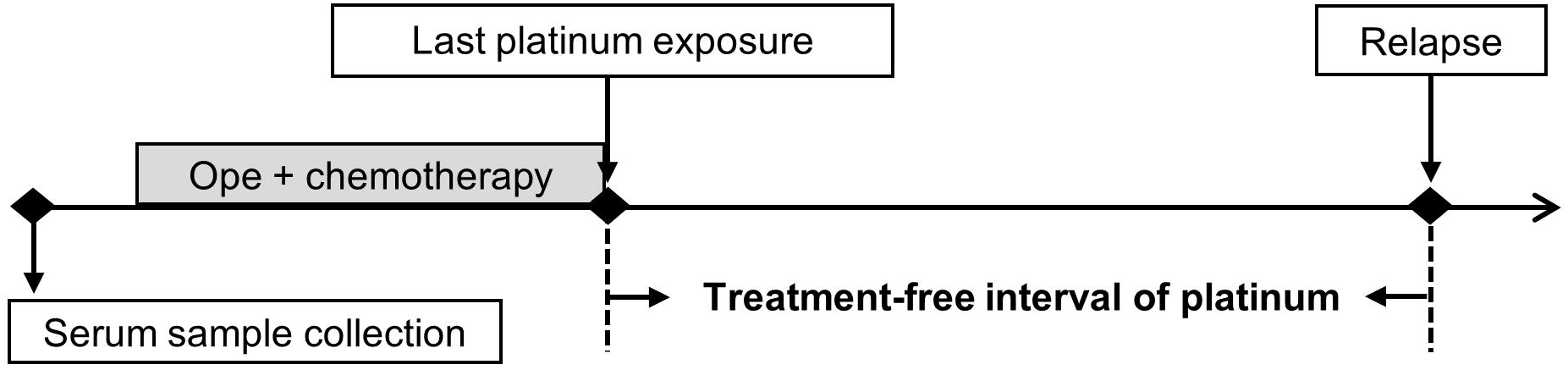
Time course of ovarian cancer treatment and definition of treatment-free interval of platinum Abbreviation: Ope, operation.

**Supplementary Figure 2.**
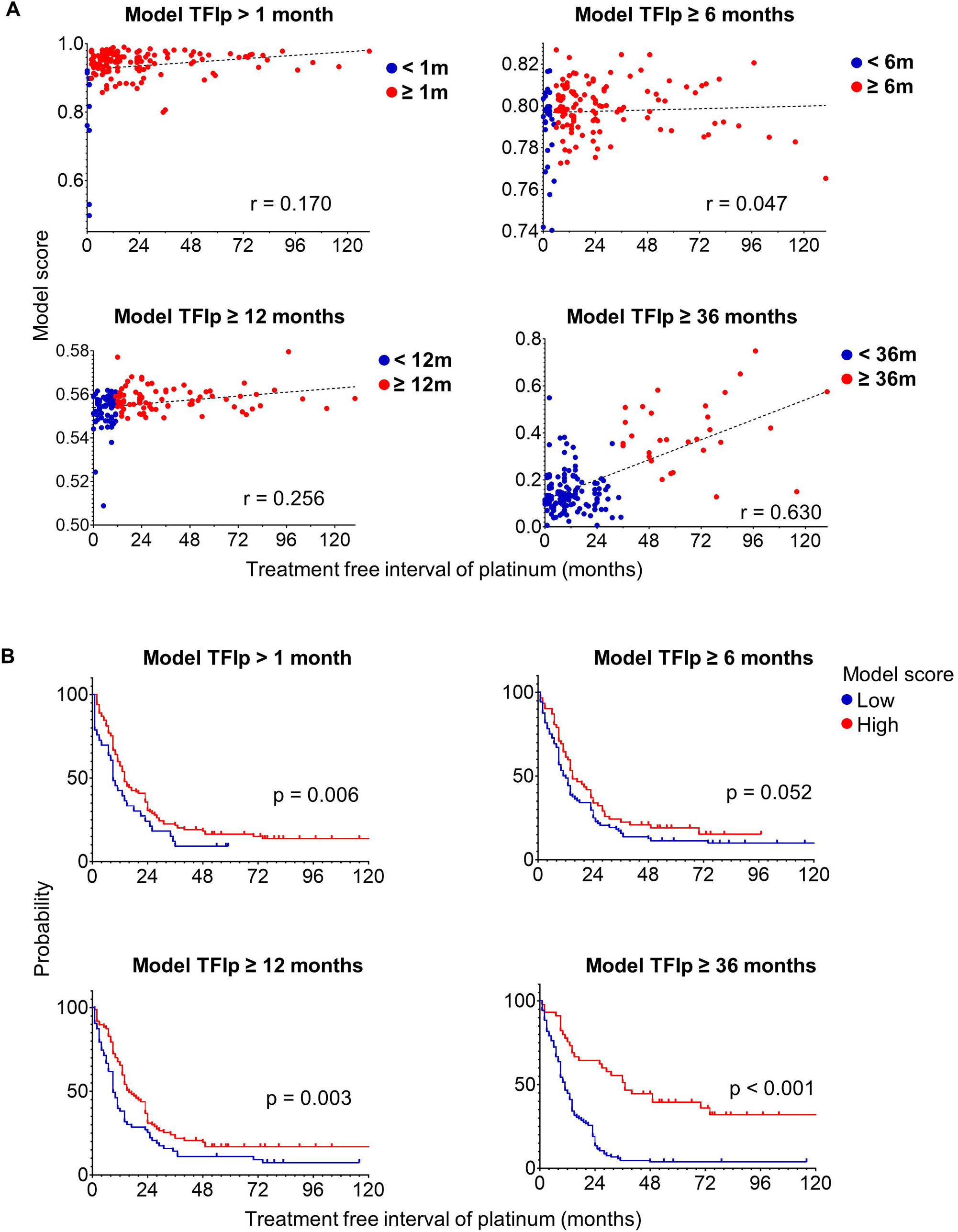
A. Dot plot for each model. Dotted lines indicate linear regression model. R values were calculated using Pearson’s correlation coefficient. B. Events over time were evaluated based on predictive models. Kaplan–Meier curves showing patients stratified into high and low groups by each model with cutoffs based on Youden index. *p* values were calculated using the generalized Wilcoxon test

**Supplementary Table 1.**
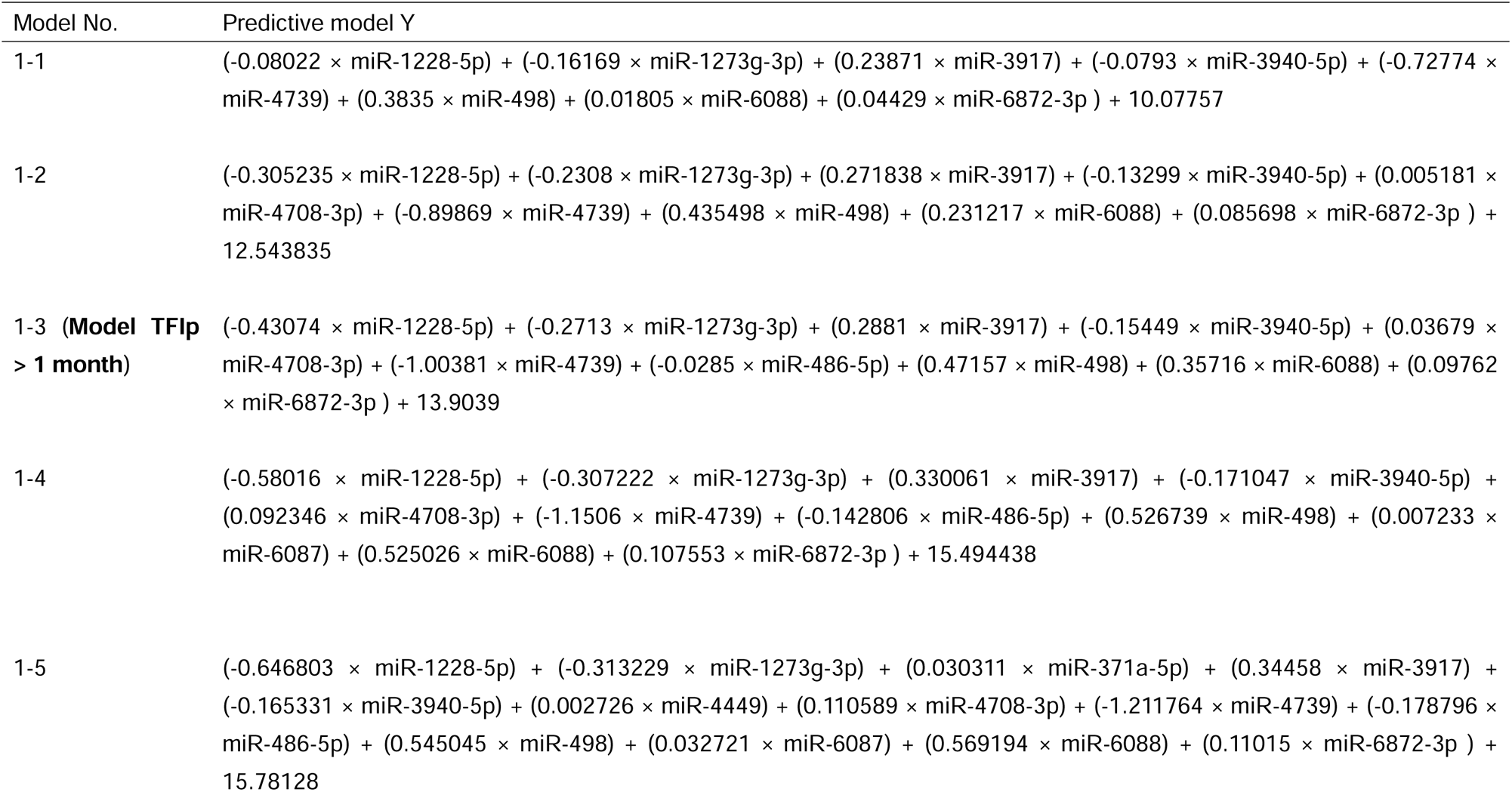
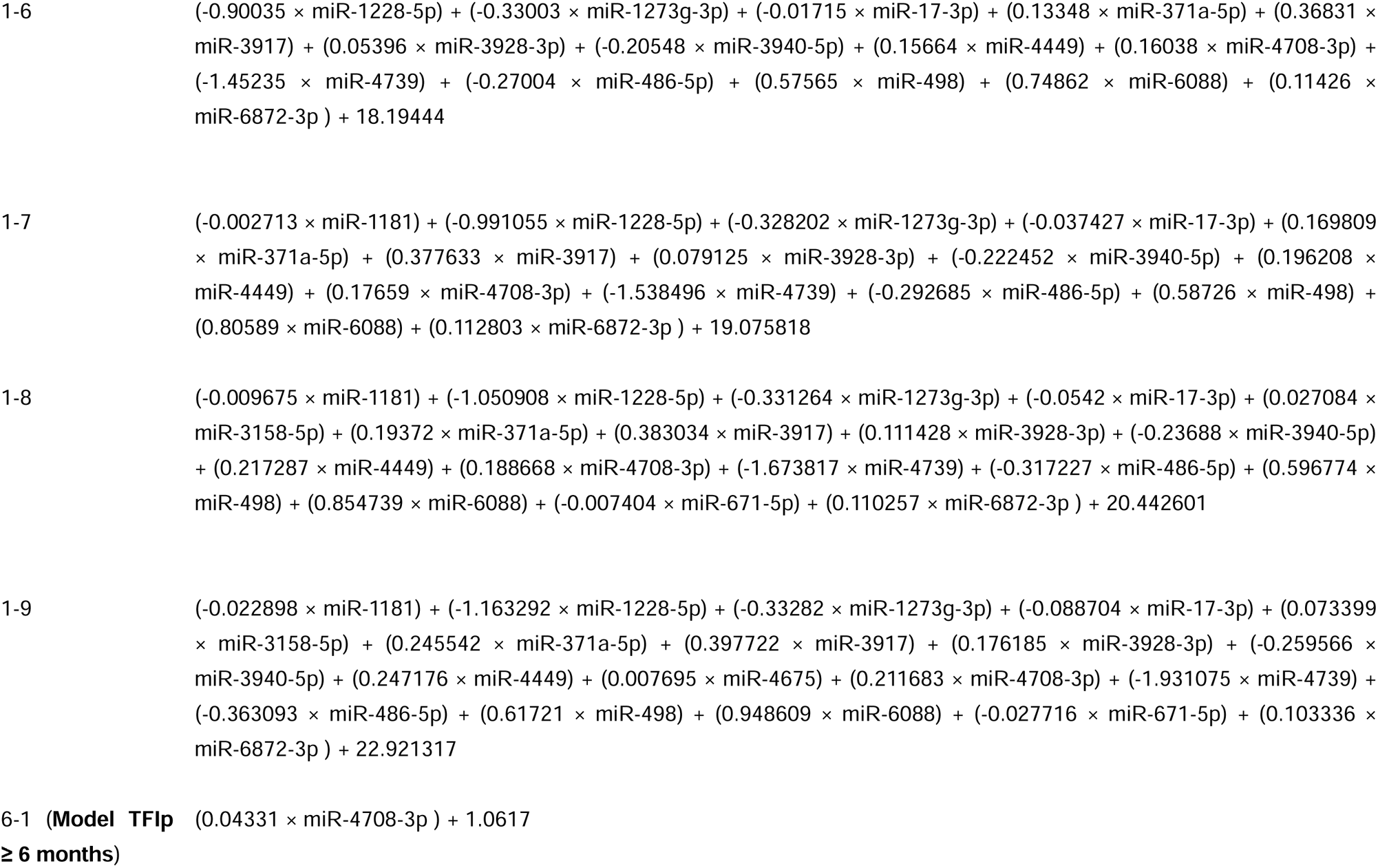
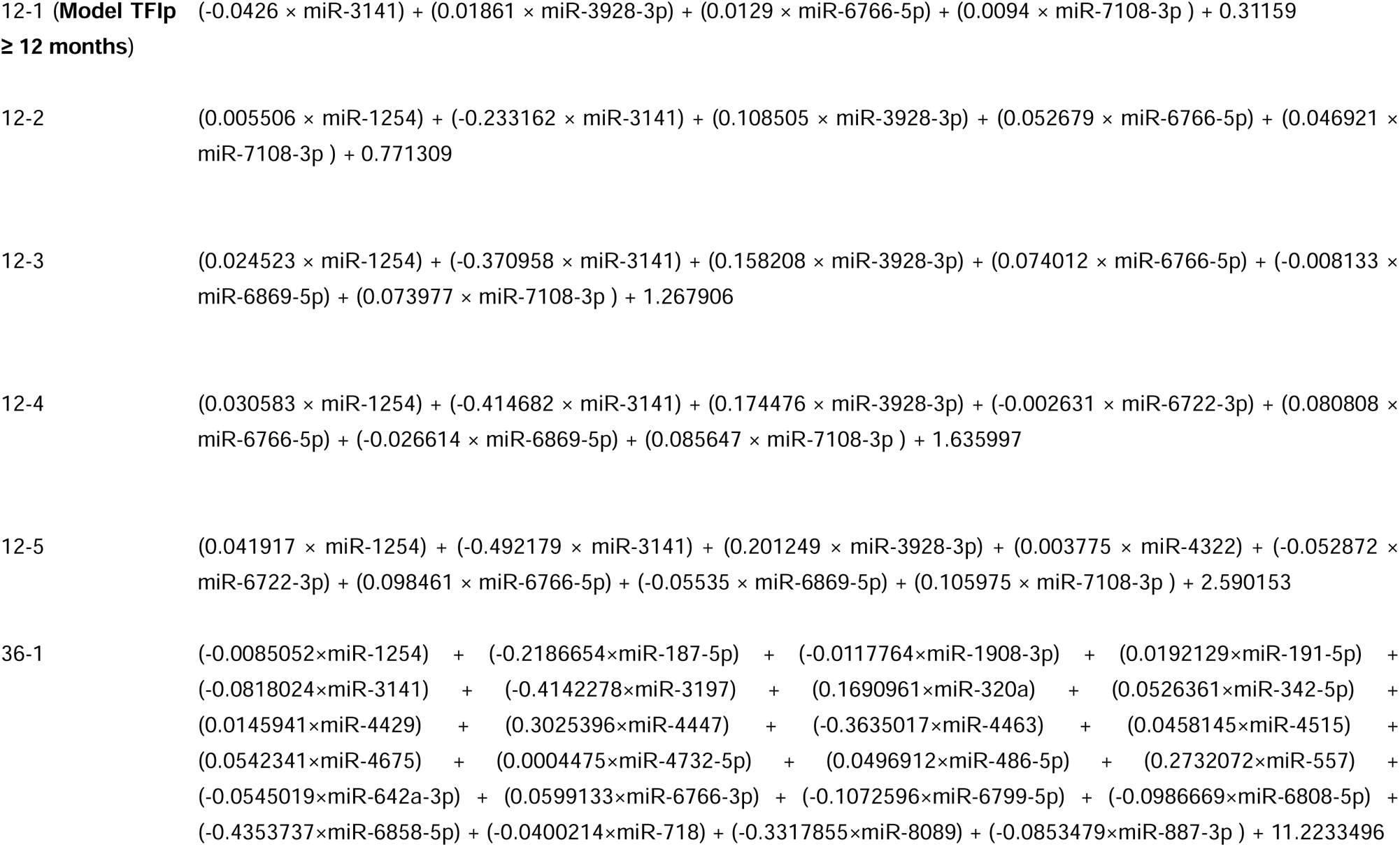
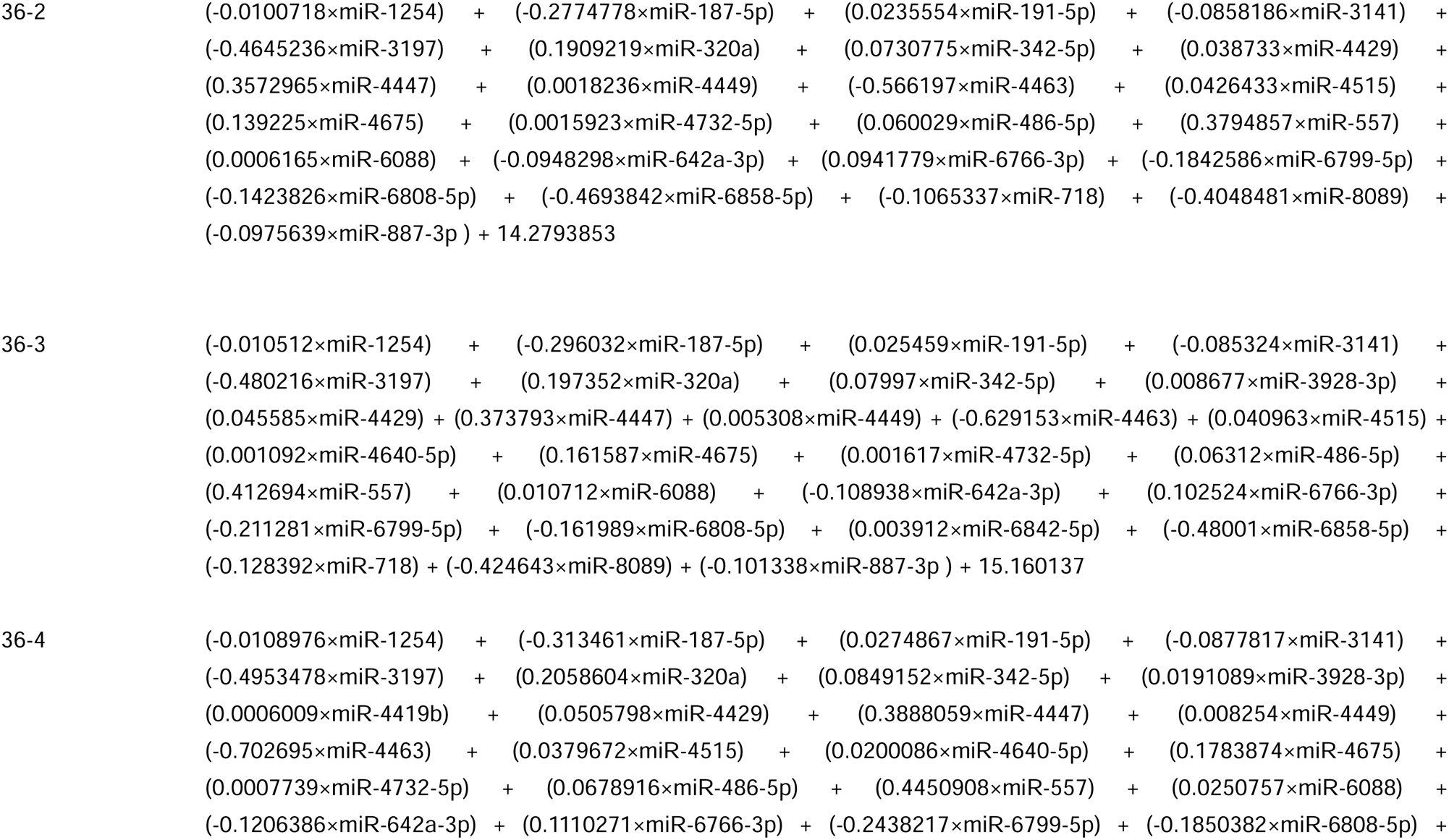
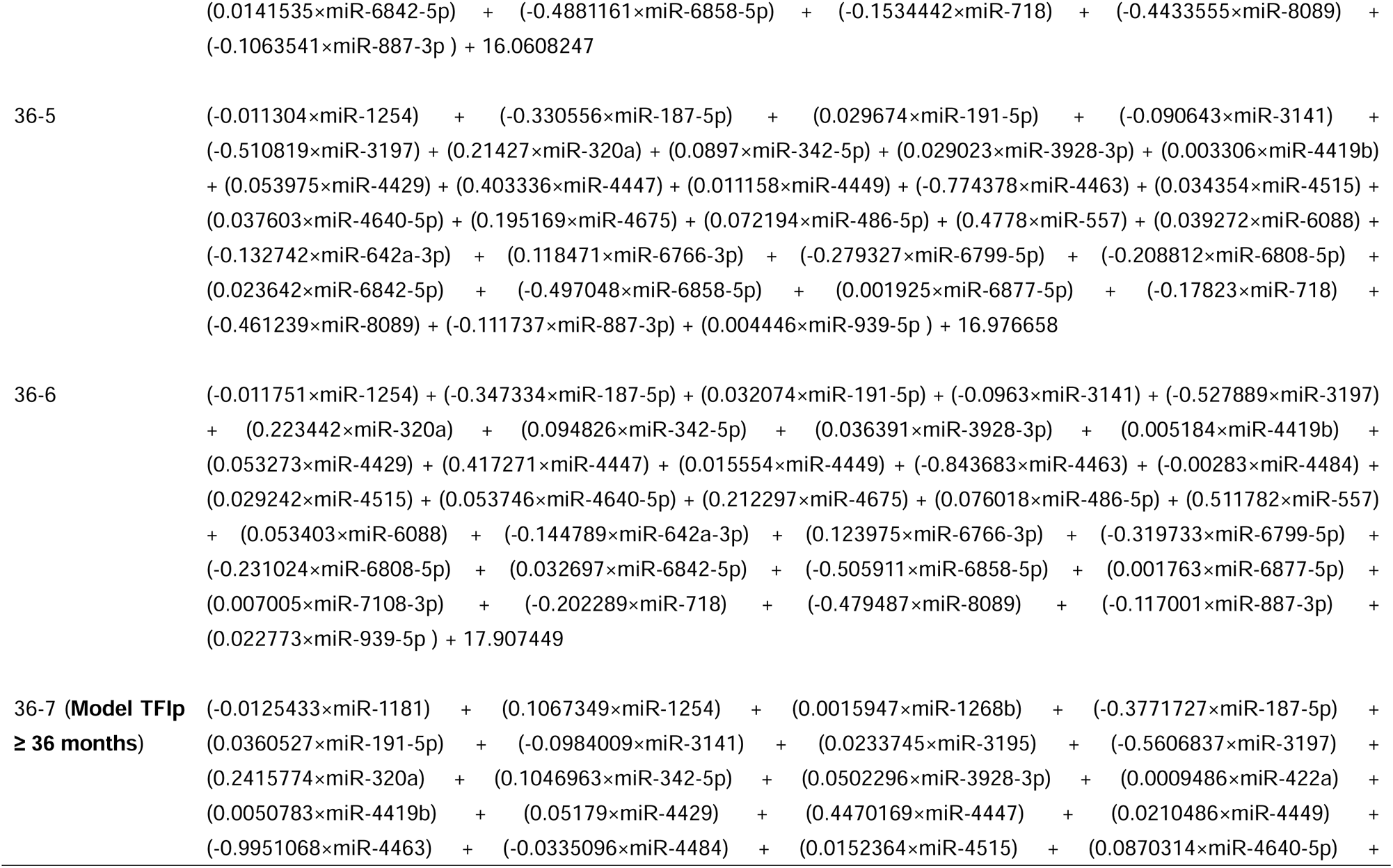
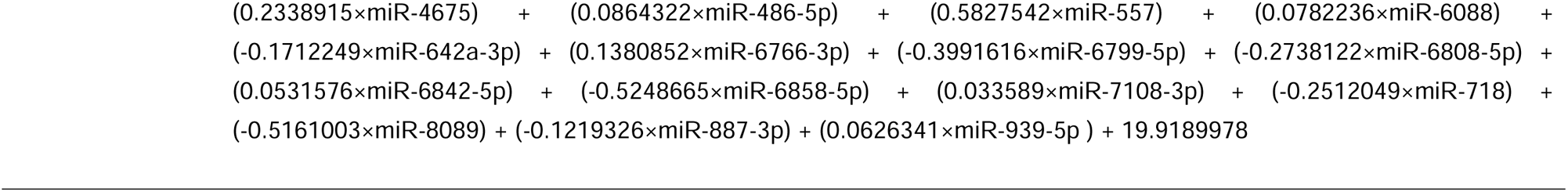
Detail of predictive model.

**Supplementary Table 2.** AUC for all subgroups based on each model.

| Model No. | Predict TFIp |  |  |  |
| --- | --- | --- | --- | --- |
|  | > 1 month | ≥ 6 months | ≥ 12 months | ≥ 36 months |
| Model TFIp > 1 month | **0.944** | **0.663** | **0.603** | **0.618** |
| Model TFIp ≥ 6 months | **0.631** | **0.637** | **0.570** | **0.563** |
| Model TFIp ≥ 12 months | **0.757** | **0.698** | **0.705** | **0.659** |
| Model TFIp ≥ 36 months | **0.624** | **0.641** | **0.645** | **0.938** |

Note: Data are shown as AUC unless otherwise noted.

## References

[1] Sung H, Ferlay J, Siegel RL, Laversanne M, Soerjomataram I, Jemal A, et al. Global Cancer Statistics 2020: GLOBOCAN Estimates of Incidence and Mortality Worldwide for 36 Cancers in 185 Countries. CA Cancer J Clin. 2021;71:209–49.

[2] Lheureux S, Gourley C, Vergote I, Oza AM. Epithelial ovarian cancer. Lancet. 2019;393:1240–53.

[3] Reid BM, Permuth JB, Sellers TA. Epidemiology of ovarian cancer: a review. Cancer Biol Med. 2017;14:9–32.

[4] Armstrong DK, Alvarez RD, Bakkum-Gamez JN, Barroilhet L, Behbakht K, Berchuck A, et al. Ovarian Cancer, Version 2.2020, NCCN Clinical Practice Guidelines in Oncology. J Natl Compr Canc Netw. 2021;19:191–226.

[5] Pujade-Lauraine E, Selle F, Scambia G, Asselain B, Marme F, Lindemann K, et al. Maintenance olaparib rechallenge in patients with platinum-sensitive relapsed ovarian cancer previously treated with a PARP inhibitor (OReO/ENGOT-ov38): a phase IIIb trial. Ann Oncol. 2023.

[6] Takamatsu S, Nakai H, Yamaguchi K, Hamanishi J, Mandai M, Matsumura N. Time-Dependent Changes in Risk of Progression During Use of Bevacizumab for Ovarian Cancer. JAMA Netw Open. 2023;6:e2326834.

[7] Pujade-Lauraine E, Alexandre J. Update of randomized trials in recurrent disease. Ann Oncol. 2011;22 Suppl 8:viii61–viii4.

[8] Pujade-Lauraine E, Hilpert F, Weber B, Reuss A, Poveda A, Kristensen G, et al. Bevacizumab combined with chemotherapy for platinum-resistant recurrent ovarian cancer: The AURELIA open-label randomized phase III trial. J Clin Oncol. 2014;32:1302–8.

[9] Arend RC, Monk BJ, Shapira-Frommer R, Haggerty AF, Alvarez EA, Amit A, et al. Ofranergene Obadenovec (Ofra-Vec, VB-111) With Weekly Paclitaxel for Platinum-Resistant Ovarian Cancer: Randomized Controlled Phase III Trial (OVAL Study/GOG 3018). J Clin Oncol. 2023:JCO2202915.

[10] van der Burg ME, Vergote I, Onstenk W, Boere IA, Leunen K, van Montfort CA, et al. Long-term results of weekly paclitaxel carboplatin induction therapy: an effective and well-tolerated treatment in patients with platinum-resistant ovarian cancer. Eur J Cancer. 2013;49:1254–63.

[11] Lortholary A, Largillier R, Weber B, Gladieff L, Alexandre J, Durando X, et al. Weekly paclitaxel as a single agent or in combination with carboplatin or weekly topotecan in patients with resistant ovarian cancer: the CARTAXHY randomized phase II trial from Groupe d’Investigateurs Nationaux pour l’Etude des Cancers Ovariens (GINECO). Ann Oncol. 2012;23:346–52.

[12] Ambros V. The functions of animal microRNAs. Nature. 2004;431:350–5.

[13] Esquela-Kerscher A, Slack FJ. Oncomirs - microRNAs with a role in cancer. Nat Rev Cancer. 2006;6:259–69.

[14] Valadi H, Ekstrom K, Bossios A, Sjostrand M, Lee JJ, Lotvall JO. Exosome-mediated transfer of mRNAs and microRNAs is a novel mechanism of genetic exchange between cells. Nat Cell Biol. 2007;9:654–9.

[15] Kosaka N, Iguchi H, Ochiya T. Circulating microRNA in body fluid: a new potential biomarker for cancer diagnosis and prognosis. Cancer Sci. 2010;101:2087–92.

[16] Yokoi A, Ochiya T. Exosomes and extracellular vesicles: Rethinking the essential values in cancer biology. Semin Cancer Biol. 2021;74:79–91.

[17] Yokoi A, Matsuzaki J, Yamamoto Y, Yoneoka Y, Takahashi K, Shimizu H, et al. Integrated extracellular microRNA profiling for ovarian cancer screening. Nat Commun. 2018;9:4319.

[18] Yoshida K, Yokoi A, Matsuzaki J, Kato T, Ochiya T, Kajiyama H, et al. Extracellular microRNA profiling for prognostic prediction in patients with high-grade serous ovarian carcinoma. Cancer Sci. 2021;112:4977–86.

[19] Suzuki K, Yokoi A, Yoshida K, Kato T, Ochiya T, Yamamoto Y, et al. Preoperative serum microRNAs as potential prognostic biomarkers in ovarian clear cell carcinoma. J Gynecol Oncol. 2023;34:e34.

[20] Okuno K, Kandimalla R, Mendiola M, Balaguer F, Bujanda L, Fernandez-Martos C, et al. A microRNA signature for risk-stratification and response prediction to FOLFOX-based adjuvant therapy in stage II and III colorectal cancer. Mol Cancer. 2023;22:13.

[21] Wu A, Yen R, Grasedieck S, Lin H, Nakamoto H, Forrest DL, et al. Identification of multivariable microRNA and clinical biomarker panels to predict imatinib response in chronic myeloid leukemia at diagnosis. Leukemia. 2023.

[22] Kozomara A, Griffiths-Jones S. miRBase: annotating high confidence microRNAs using deep sequencing data. Nucleic Acids Res. 2014;42:D68–73.

[23] Yu X, Zhang X, Wang G, Wang B, Ding Y, Zhao J, et al. miR-206 as a prognostic and sensitivity biomarker for platinum chemotherapy in epithelial ovarian cancer. Cancer Cell Int. 2020;20:534.

[24] Compadre AJ, van Biljon LN, Valentine MC, Llop-Guevara A, Graham E, Fashemi B, et al. RAD51 Foci as a Biomarker Predictive of Platinum Chemotherapy Response in Ovarian Cancer. Clin Cancer Res. 2023;29:2466–79.

[25] Ishida S, McCormick F, Smith-McCune K, Hanahan D. Enhancing tumor-specific uptake of the anticancer drug cisplatin with a copper chelator. Cancer Cell. 2010;17:574–83.

[26] Rizzo S, Hersey JM, Mellor P, Dai W, Santos-Silva A, Liber D, et al. Ovarian cancer stem cell-like side populations are enriched following chemotherapy and overexpress EZH2. Mol Cancer Ther. 2011;10:325–35.

[27] Latifi A, Abubaker K, Castrechini N, Ward AC, Liongue C, Dobill F, et al. Cisplatin treatment of primary and metastatic epithelial ovarian carcinomas generates residual cells with mesenchymal stem cell-like profile. J Cell Biochem. 2011;112:2850–64.

[28] Belanger F, Fortier E, Dube M, Lemay JF, Buisson R, Masson JY, et al. Replication Protein A Availability during DNA Replication Stress Is a Major Determinant of Cisplatin Resistance in Ovarian Cancer Cells. Cancer Res. 2018;78:5561–73.

[29] Zheng D, Huang X, Peng J, Zhuang Y, Li Y, Qu J, et al. CircMYOF triggers progression and facilitates glycolysis via the VEGFA/PI3K/AKT axis by absorbing miR-4739 in pancreatic ductal adenocarcinoma. Cell Death Discov. 2021;7:362.

[30] Chen Y, Wang X. miRDB: an online database for prediction of functional microRNA targets. Nucleic Acids Res. 2020;48:D127–D31.

[31] Khosravi N, Caetano MS, Cumpian AM, Unver N, De la Garza Ramos C, Noble O, et al. IL22 Promotes Kras-Mutant Lung Cancer by Induction of a Protumor Immune Response and Protection of Stemness Properties. Cancer Immunol Res. 2018;6:788–97.

[32] Wang S, Yao Y, Yao M, Fu P, Wang W. Interleukin-22 promotes triple negative breast cancer cells migration and paclitaxel resistance through JAK-STAT3/MAPKs/AKT signaling pathways. Biochem Biophys Res Commun. 2018;503:1605–9.

[33] Wu D, Zhou J, Tan M, Zhou Y. LINC01116 regulates proliferation, migration, and apoptosis of keloid fibroblasts by the TGF-beta1/SMAD3 signaling via targeting miR-3141. Anal Biochem. 2021;627:114249.

[34] Zhu H, Gu X, Xia L, Zhou Y, Bouamar H, Yang J, et al. A Novel TGFbeta Trap Blocks Chemotherapeutics-Induced TGFbeta1 Signaling and Enhances Their Anticancer Activity in Gynecologic Cancers. Clin Cancer Res. 2018;24:2780–93.

[35] Gagno S, Poletto E, Bartoletti M, Quartuccio L, Romualdi C, Garziera M, et al. A TGF-beta associated genetic score to define prognosis and platinum sensitivity in advanced epithelial ovarian cancer. Gynecol Oncol. 2020;156:233–42.

[36] Ghezelayagh TS, Pennington KP, Norquist BM, Khasnavis N, Radke MR, Kilgore MR, et al. Characterizing TP53 mutations in ovarian carcinomas with and without concurrent BRCA1 or BRCA2 mutations. Gynecol Oncol. 2021;160:786–92.

[37] You B, Robelin P, Tod M, Louvet C, Lotz JP, Abadie-Lacourtoisie S, et al. CA-125 ELIMination Rate Constant K (KELIM) Is a Marker of Chemosensitivity in Patients with Ovarian Cancer: Results from the Phase II CHIVA Trial. Clin Cancer Res. 2020;26:4625–32.

[38] Lindholm EM, Ragle Aure M, Haugen MH, Kleivi Sahlberg K, Kristensen VN, Nebdal D, et al. miRNA expression changes during the course of neoadjuvant bevacizumab and chemotherapy treatment in breast cancer. Mol Oncol. 2019;13:2278–96.

[39] Choi YE, Meghani K, Brault ME, Leclerc L, He YJ, Day TA, et al. Platinum and PARP Inhibitor Resistance Due to Overexpression of MicroRNA-622 in BRCA1-Mutant Ovarian Cancer. Cell Rep. 2016;14:429–39.

